# Hypnosis versus relaxation for anxiety in COPD pulmonary rehabilitation: a cluster-randomised trial

**DOI:** 10.64898/2026.07.20.26358482

**Authors:** Adelina Ghergan, Bertrand Herer, François Larue, Delphine Sambourg, Isabelle Segundo, Yolaine Bocahu, Cécile Moulin, Agathe Delignières, Thomas Similowski, Capucine Morelot-Panzini, Hernán Anlló

## Abstract

**Background:** Anxiety affects 22–56% of patients with chronic obstructive pulmonary disease and predicts exacerbations, readmissions, and mortality. Mind-body adjuncts to pulmonary rehabilitation are recommended, but trials have lacked comparators isolating technique-specific effects and left determinants of individual benefit unexamined. We tested whether adjunctive medical hypnosis reduces anxiety more than matched structured relaxation.

**Methods:** Cluster-randomised, active-comparator trial (ClinicalTrials.gov NCT04868357) with patients undergoing pulmonary rehabilitation for chronic obstructive pulmonary disease. Consecutive 4-week inpatient cohorts were randomised 1:1 to adjunctive hypnosis or structured relaxation matched on session length, practitioner pool, and framing. Primary outcome was the six-item State-Trait Anxiety Inventory at week 4 (intention to treat). Complementary multidimensional metrics of anxiety were obtained for robustness. Exploratory analyses tested outcome moderation by baseline sensory-affective dyspnoea burden.

**Results:** Of 80 participants (24 clusters), 79 were analysed (hypnosis 36, relaxation 43). Hypnosis did not differ from relaxation on the primary outcome (+0.95 points; 95% credible interval −5.85 to 7.42; posterior probability of clinically important hypnosis advantage 0.12) or its complementary metrics. Anxiety improved in both arms (pooled −5.72 points; −9.35 to −2.00). Exploratory analyses showed that the hypnosis advantage rose with baseline sensory burden, and fell with affective burden, reaching −9.26 points at a high-sensory, low-affective profile (−23.37 to 4.71; posterior probability of clinically important hypnosis advantage 0.81). No deaths or serious adverse events occurred.

**Conclusions:** Anxiety improved with no advantage of hypnosis over matched relaxation. Exploratory analysis suggests greater hypnosis benefit in patients with predominantly sensory dyspnoea burden, proposing a candidate phenotype for stratified trials.

## Introduction

Anxiety is among the most prevalent and least adequately managed comorbidities of chronic obstructive pulmonary disease (COPD), affecting 22% to 56% of patients across populations and disease stages (1–5). It is associated with worse dyspnoea, reduced exercise tolerance, greater health care utilisation, and higher risks of 30-day readmission, exacerbation, and death, independent of disease severity (1, 2). Pharmacological management of dyspnoea-associated anxiety in COPD has shown mixed efficacy (3). The 2026 Global Initiative for Chronic Obstructive Lung Disease strategy recommends combining pulmonary rehabilitation (PR) with cognitive-behavioural therapy and mind-body interventions (e.g., mindfulness-based therapy, yoga, relaxation) to alleviate anxiety and depressive symptoms (3, 4). However, the benefit of these interventions is heterogeneous, with a substantial subset of patients deriving limited improvement (5), calling for a better understanding of what determines intervention success.

Mind-body adjuncts to PR are increasingly studied, but the evidence base is limited by comparator choice. Most trials of cognitive-behavioural therapy (6), mindfulness-based interventions (7), and hypnosis (8) in COPD have used waitlist or treatment-as-usual comparators that conflate intervention-specific effects with the nonspecific benefits of any structured psychological contact embedded in PR. The present trial addresses this gap with an active comparator matched on these nonspecific components, with a preregistered multi-instrument approach to measure anxiety across multiple clinically relevant constructs (STAI-6, MDP, HADS-A). The trial focuses on medical hypnosis, based on the strength of two precedents (9, 10). First, a single 15-minute session of medical hypnosis delivered to inpatients with severe COPD produced an immediate and clinically meaningful reduction in state anxiety (9), establishing proof of concept in this population. Second, mechanistic work in healthy volunteers under inspiratory threshold loading and hypercapnia with restrained ventilation showed that hypnosis attenuates both the sensory and affective dimensions of experimental dyspnoea through combined effects on breathing drive and cortical engagement (10). Together, these findings indicate that hypnosis acts on both anxiety and dyspnoea perception and may be a viable low-cost adjunct for anxiety management in COPD. Whether this anxiolytic response is reliable in clinical practice, and whether it is uniform across patients or different expressions of dyspnoea, remained unanswered.

In this trial, we evaluated the efficacy of medical hypnosis as an adjunct to inpatient PR for anxiety in COPD, using structured relaxation as an active comparator matched on session length, structure, practitioner pool, and self-management framing. By holding these nonspecific components constant, we aimed to isolate the cognitive-control components more specific to hypnosis, namely sensory suggestion and interoceptive redirection (11, 12). We hypothesised that both arms would reduce anxiety and, as the trial’s confirmatory objective, that adjunctive hypnosis would reduce anxiety more than structured relaxation. Separately, in an exploratory post hoc moderator analysis, we examined whether the hypnotic advantage varied with baseline sensory and affective dyspnoea burden.

## Methods

Additional details are provided in the supplementary material.

### Study design and participants

HYPNOBPCO_2 (NCT04868357; www.clinicaltrials.gov) (13) was a phase 2, single-centre, parallel-group, cluster-randomised, active-comparator superiority trial of medical hypnosis versus structured relaxation as adjuncts to a 4-week inpatient pulmonary rehabilitation (PR) programme for COPD at Centre Hospitalier de Bligny (CHB; Briis-sous-Forges, France). The protocol was approved by the Comité de Protection des Personnes Île-de-France 1 (2019-A02016-51) and has been published (13). All participants provided written informed consent. Reporting follows the Consolidated Standards of Reporting Trials (CONSORT) 2025 statement and its cluster-randomised and non-pharmacological treatment extensions.

All patients admitted to PR between September 27, 2021, and January 31, 2024, were screened consecutively. Eligible participants were adults older than 30 years with COPD confirmed by spirometry (forced expiratory volume in 1 second/forced vital capacity <0.70), a modified Medical Research Council dyspnoea grade of 2 or higher, and at least 10 pack-years of tobacco exposure. Exclusion criteria (supplementary material, S2.2) included pregnancy, severe cardiac insufficiency or pulmonary arterial hypertension, active malignancy, hypercapnic encephalopathy, deafness, anaemia, psychotic illness, and clinically significant cognitive impairment. Sex was recorded as sex assigned at birth.

### Randomisation and masking

The unit of randomisation was the consecutive PR cohort (cluster). Patients were assigned to cohorts on clinical grounds, and each cohort was constituted before it was randomised, so neither allocation nor its foreknowledge could influence cohort composition. Allocation was paired at the cluster level using sealed, sequentially numbered, opaque envelopes prepared by a sponsor agent independent of the trial team: 12 pairs, each containing one hypnosis and one relaxation allocation, yielding a 1:1 ratio across 24 clusters. Envelopes were drawn immediately before each cohort began by a hospital representative independent of clinical care (S3). Participants and practitioners were necessarily unmasked. Outcome data collectors worked from datasets coded by envelope number, telephone interviewers were masked, and the trial statistician remained masked until the primary analyses were complete (S4).

### Interventions

All participants received the centre’s standard 4-week residential PR programme: approximately 30 hours per week of cycle endurance training at 75% of peak work rate, strength training, COPD self-management workshops, dyspnoea-desensitisation workshops, and speech and singing therapy, plus needs-based psychological counselling and smoking-cessation support, identical in both arms (S5.1).

Participants in both arms also attended two 45-minute group sessions, on the Wednesdays of weeks 1 and 3. Medical hypnosis sessions followed a standardised script comprising a hypnotic induction, suggestions of relaxation and of "air entering effortlessly into the lungs," nature-themed metaphors, and an explicit prescription to practice self-hypnosis for anxiety and dyspnoea throughout PR. Structured relaxation sessions followed a standardised script of identical duration, structure, and prescription, comprising six dynamic relaxation exercises focused on proprioception, calmness, and breath regulation. Scripts were not tailored (Appendices 3 and 4). Both interventions were delivered by the same three practitioners (A.D., C.M., I.S.; Association Française d’Hypnose–certified, ≥4 years’ experience), rotated by duty roster so that practitioner was fully crossed with allocation. Fidelity rested on the verbatim scripts, practitioner certification and rotation, and a pass/fail delivery check performed once per cohort by the investigators leading clinical conduct (A.G. and B.H.; all passed); sessions were not recorded or rated at component level (S5.6).

### Outcomes

The primary outcome was the six-item State-Trait Anxiety Inventory (STAI-6) score (14) at week 4, adjusted for baseline. Prespecified confirmatory measures of the same anxiety construct were the Hospital Anxiety and Depression Scale anxiety subscale (HADS-A) (15) and the affective subscales of the Multidimensional Dyspnea Profile (MDP) (16): MDP-A1, the MDP-A2 sum, and its anxiety and fear items, treated as one coordinated operationalisation without multiplicity correction (17, 18). Secondary outcomes were the MDP sensory subscales (MDP-SQ sum, air hunger, and work/effort), the COPD Assessment Test (CAT) (19), and 6-minute walk distance (20); the HADS depression subscale was another registered outcome. All were assessed at weeks 1 and 4 (except STAI-6, which was assessed weekly). Prespecified telephone follow-up of STAI-6 and CAT at 1, 2, and 3 months after discharge was analysed descriptively because of substantial differential attrition (S8.11 and Table S6). Adverse events were solicited at every study visit and recorded with their severity, seriousness, and relationship to the intervention (S10). Because these are patient-reported outcomes, the outcome source was necessarily unmasked. The registration listed the STAI-6, HADS, and MDP as co-primary outcomes; analyses follow the published protocol (13), which designated the STAI-6 as primary and the others as additional robustness metrics, a redesignation made before any analysis (S8.2) which did not alter the results of the trial in any way.

### Statistical analysis

The sample size was derived by simulation for a mixed-effects model with cluster random intercepts (13): 10 clusters of five participants per arm (n=100) gave 83% power (95% confidence interval, 80–86%) at a two-sided α of 0.05 to detect a 10% (approximately 4.7-point) additional STAI-6 reduction with hypnosis, assuming a 24% reduction with hypnosis, as in HYPNOBPCO_1 (9), and 14% with relaxation from a baseline of 47 points. The target was set at 120 to allow for 20% attrition. Given the novelty of the approach, no empirical pre-existing intracluster correlation coefficient was available at simulation time.

All analyses followed the intention-to-treat principle and included all randomised participants except those who withdrew consent. Because the programme was residential, the intention-to-treat and per-protocol sets were identical by design. Missing items were handled by multiple imputation by chained equations (M = 100, item level) with predictive mean matching under a missing-at-random assumption; differential attrition at month 3 was addressed by a tipping-point sensitivity analysis (S8.3 and S8.11).

The primary analysis was a Bayesian analysis of covariance with a cluster-level random intercept, which adjusted for the baseline value of the outcome, baseline CAT, baseline 6-minute walk distance, age, and sex, fitted across the imputed datasets with weakly informative priors whose central 95% interval spanned several minimal clinically important differences (MCIDs) on each scale (S8.4). For every outcome we report the posterior median between-arm contrast (hypnosis minus relaxation), its 95% highest-density interval (HDI), and the posterior probability that the contrast exceeded the MCID (STAI-6, 3 points, a precedent-based threshold in the absence of a COPD-specific MCID (S8.9); HADS-A, 1.5; MDP items, 1; MDP sums, 3; CAT, 2; 6-minute walk distance, 30 m). The MDP thresholds are conservative and author-defined, as no validated MCID exists. They are used descriptively only. No Bayesian decision rule was prespecified; posterior probabilities are descriptive summaries, not success criteria. The intracluster correlation coefficient was derived from the posterior variance components (21–23). This approach replaced the registered constrained longitudinal model as the primary analysis to account for an unexpected baseline imbalance between arms, observed on arm-coded data before unmasking. The registered model is retained nonetheless, as a secondary analysis (S8.5). Of note, the change was found to be conservative with respect to the trial hypothesis. Analysis of covariance conditions explicitly on each outcome’s baseline value, and because the hypnosis arm entered the trial with lower anxiety, adjusting for baseline removes a head start that the registered model, which estimates each arm’s trajectory from its observed starting point, would have credited to hypnosis (Results; S8.5).

A post hoc, exploratory moderation analysis, grounded in the sensory–affective structure of the MDP (16) and prior mechanistic work (9, 10), tested whether the hypnosis effect varied with baseline dyspnoea burden. Two standardised composite indices were formed from preregistered week-1 values: a sensory-somatic index (mean z-score of the MDP-SQ sum and CAT) and an affective index (mean z-score of the MDP-A2 sum, HADS-A, and STAI-6), so that moderator and outcome roles were separated by time point (S7). Bayesian analyses of covariance with continuous moderator-by-intervention interactions were fitted to the primary and confirmatory anxiety outcomes, and between-arm contrasts were reported at +1 SD on the sensory and −1 SD on the affective index, as an illustrative point on a continuum. As part of this analytical strategy, HADS-A, which is not anchored to the respiratory experience, was designated post hoc as a within-construct negative control on which no gradient was expected. For robustness, the moderation was also re-estimated with baseline STAI-6 and MDP-A2 sum removed from the index subsequently, so that no outcome moderated itself, and with the sensory index restricted to the MDP-SQ sum, removing the CAT’s non-sensory items (Table 4).

Five refinements to the registered plan were specified before unmasking (S8.2): the replacement of the constrained longitudinal model by baseline-adjusted analysis of covariance as the primary analysis, because of a baseline imbalance observed on arm-coded data as explained above; the addition of multiple imputation; analysis of the MDP at its sensory and affective subscales rather than just as a composite; full posterior estimation, expanding on the protocol’s original Bayesian analysis strategy; and the descriptive treatment of the month 1–3 assessments given differential attrition. Analyses were performed in R version 4.6.0.

## Results

### Participants

Between September 27, 2021, and January 31, 2024, 188 patients admitted to PR in 25 consecutive cohorts were screened, 65 were not enrolled, and 123 consented (Figure 1). Following the developing COVID-19 situation at the time, 43 consented patients were reclassified at the admission diagnostic assessment to post-COVID-19 condition or other non-COPD aetiologies (S2.3). One cohort, left with no eligible participant, was not randomised. Reclassification and smaller pandemic-era cohorts left 80 participants randomised in 24 clusters, below the 100 required: 36 to medical hypnosis and 44 to structured relaxation (12 clusters each; cluster size 1–7). One relaxation participant withdrew consent after allocation, before any data were collected, leaving 79 analysed. All 79 attended both sessions and completed the residential programme; 72 provided the week-4 primary outcome (Table S8). The last follow-up assessment was on May 24, 2024.

**Figure 1.**
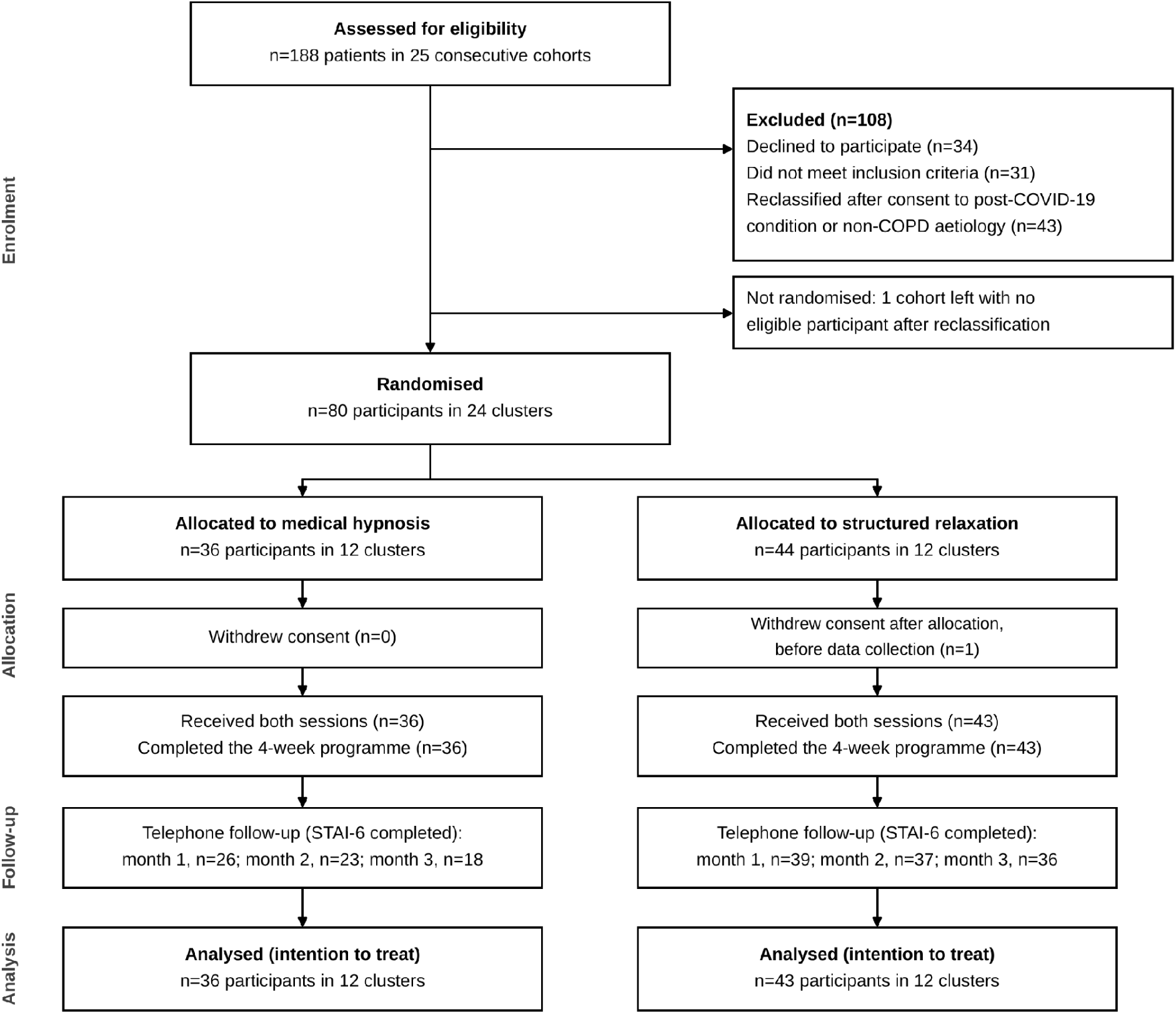
Participant flow (CONSORT 2025 flow diagram with the cluster-randomised extension). Patients were screened consecutively on admission to pulmonary rehabilitation; cohorts were randomised as clusters after they were constituted. Reclassification refers to diagnostic reclassification to post-COVID-19 condition or other non-COPD aetiologies at the admission assessment, before randomisation.

Table 1 shows baseline characteristics. Compared with the relaxation arm, the hypnosis arm had lower baseline STAI-6 (standardised mean difference, −0.37), higher MDP-A2 sum (+0.41) and fear item (+0.30), and higher arterial carbon dioxide tension (PaCO₂; +0.62). PaCO₂ lay outside the prespecified covariate set and was examined in a sensitivity analysis.

**Table 1.** Baseline characteristics by trial arm

| **Characteristic** | **Medical hypnosis<br>(n=36)** | **Structured relaxation<br>(n=43)** | **n available<br>(hypnosis/relaxation)** |
|---|---|---|---|
| **Demographics** |  |  |  |
| Age, yr, mean (SD)* | 64.9 (8.3) | 67.4 (9.1) | 36/43 |
| Sex, female, n (%)* | 20 (56) | 23 (53) | 36/43 |
| **Anthropometrics** |  |  |  |
| BMI, kg/m<sup>2</sup>, mean (SD) | 25.4 (5.5) | 24.8 (5.8) | 34/40 |
| **Anxiety and dyspnoea (baseline questionnaires), mean (SD)** |  |  |  |
| STAI-6 | 37.0 (13.4) | 42.5 (15.7) | 35/32 |
| HADS-A | 8.6 (3.5) | 9.3 (4.5) | 31/41 |
| MDP-A1 | 5.66 (2.51) | 5.53 (2.08) | 32/36 |
| MDP-A2 sum (emotional) | 18.0 (13.4) | 12.7 (12.3) | 32/33 |
| MDP-A2 anxiety item | 4.7 (3.5) | 3.9 (3.6) | 33/35 |
| MDP-A2 fear item | 3.5 (3.7) | 2.4 (3.5) | 32/36 |
| MDP-SQ sum (sensory) | 22.8 (11.8) | 20.8 (12.0) | 30/33 |
| MDP-SQ2 air hunger | 5.8 (2.9) | 4.9 (2.8) | 33/34 |
| MDP-SQ2 work/effort | 3.7 (3.1) | 4.3 (2.8) | 33/34 |
| CAT total* | 21.8 (6.4) | 21.3 (5.5) | 36/42 |
| **Spirometry and gas exchange, mean (SD)** |  |  |  |
| FEV<sub>1</sub>, % predicted | 43.4 (13.4) | 44.2 (15.6) | 34/40 |
| FVC, % predicted | 70.1 (14.4) | 69.6 (19.1) | 34/40 |
| FEV<sub>1</sub>/FVC, % | 49.7 (11.3) | 49.3 (10.6) | 34/40 |
| PaO<sub>2</sub>, mm Hg | 72.9 (13.0) | 74.5 (9.9) | 32/40 |
| PaCO<sub>2</sub>, mm Hg | 43.3 (6.8) | 39.6 (5.0) | 33/40 |
| Arterial pH | 7.42 (0.03) | 7.43 (0.02) | 32/40 |
| **COPD severity** |  |  |  |
| GOLD spirometric stage 1/2/3/4, n | 0/10/20/4 | 1/16/14/9 | 34/40 |
| GOLD group B/E, n | 8/26 | 15/25 | 34/40 |
| mMRC dyspnoea grade 2/3/4, n | 17/18/1 | 25/18/0 | 36/43 |
| BODE index, mean (SD) | 4.2 (2.0) | 4.0 (1.8) | 34/40 |
| **Smoking history** |  |  |  |
| Pack-years, mean (SD) | 49.0 (21.6) | 47.6 (26.6) | 34/40 |
| Current/former/never smoker, n | 14/19/1 | 13/27/0 | 34/40 |
| **Comorbidity and medication** |  |  |  |
| Charlson Comorbidity Index, mean (SD) | 4.2 (2.2) | 4.3 (1.8) | 34/40 |
| Cardiovascular comorbidity, n (%) | 18 (53) | 30 (75) | 34/40 |
| Long-term oxygen therapy, n (%) | 13 (38) | 17 (42) | 34/40 |
| Long-acting β-agonist, n (%) | 33 (97) | 40 (100) | 34/40 |
| Long-acting muscarinic antagonist, n (%) | 32 (94) | 35 (88) | 34/40 |
| Inhaled corticosteroid, n (%) | 22 (65) | 27 (68) | 34/40 |
| Exacerbation in past year, n (%) | 13 (38) | 11 (28) | 34/40 |
| **Mental health and analgesia, n (%)** |  |  |  |
| Psychotropic use (history) | 10 (29) | 10 (25) | 34/40 |
| Benzodiazepine use | 8 (24) | 9 (22) | 34/40 |
| Antidepressant use | 7 (21) | 9 (22) | 34/40 |
| Pain reported at baseline | 20 (59) | 20 (50) | 34/40 |
| Analgesic use | 20 (59) | 20 (50) | 34/40 |
| **Exercise capacity, mean (SD)** |  |  |  |
| 6-MWT distance, m* | 377.2 (115.6) | 380.0 (91.6) | 36/42 |
| **Cluster structure** |  |  |  |
| Number of clusters | 12 | 12 | — |
| Cluster size, mean (SD) | 3.00 (1.6) | 3.58 (1.7) | — |
| Cluster size range | 1–6 | 1–7 | — |
| Singleton clusters, n | 3 | 1 | — |

Values are mean (SD) or n (%), computed on complete cases per variable (participants with every item of the instrument present); the n available column gives the per-arm complete-case denominator (analysed sample: 36 hypnosis, 43 relaxation), and instrument-level availability is given in Table S8 (supplementary material). Inferential analyses use the multiply imputed datasets (M = 100), whose pooled baseline means (Table 2) may differ from the complete-case values shown here (supplementary material, S8.3).
BMI: body mass index; BODE: body mass index, airflow obstruction, dyspnoea, and exercise capacity index; CAT: COPD Assessment Test; COPD: chronic obstructive pulmonary disease; FEV<sub>1</sub>: forced expiratory volume in 1 second; FVC: forced vital capacity; GOLD: Global Initiative for Chronic Obstructive Lung Disease; HADS-A: Hospital Anxiety and Depression Scale, anxiety subscale; MDP: Multidimensional Dyspnea Profile (A1: immediate perception; SQ: sensory quality items; A2: emotional response items); mMRC: modified Medical Research Council dyspnoea scale; PaCO<sub>2</sub>: arterial partial pressure of carbon dioxide; PaO<sub>2</sub>: arterial partial pressure of oxygen; STAI-6: six-item State-Trait Anxiety Inventory; 6-MWT: 6-minute walk test.
\*Prespecified covariate in the primary model.

### Primary outcome

Anxiety decreased over the programme as a whole: the pooled within-arm change in STAI-6 from week 1 to week 4 was −5.72 points (95% HDI, −9.35 to −2.00; posterior probability of exceeding the MCID, 0.93; d = −0.40; per-arm estimates in S8.13). Medical hypnosis did not differ from structured relaxation on week-4 STAI-6: adjusted mean difference, +0.95 (95% HDI, −5.85 to 7.42; standardised effect size d = 0.07), with a posterior probability of hypnosis advantage of 0.38 and of exceeding the 3-point MCID of 0.12 (Table 2). At the realised sample (intracluster correlation 0.127, design effect 1.29), the minimal detectable between-arm effect at 80% power for the primary analysis was approximately 7.5 STAI-6 points (S12); the posterior is more informative than this threshold, placing 0.62 of its mass on a clinically negligible between-arm difference and at most 0.06 on an advantage of two MCIDs or more in either direction. The registered trajectory model likewise converged to a null week-4 contrast (−1.08; 95% HDI, −7.54 to 5.47; P[hypnosis < relaxation] = 0.63); both estimates lie within the MCID.

**Table 2.**
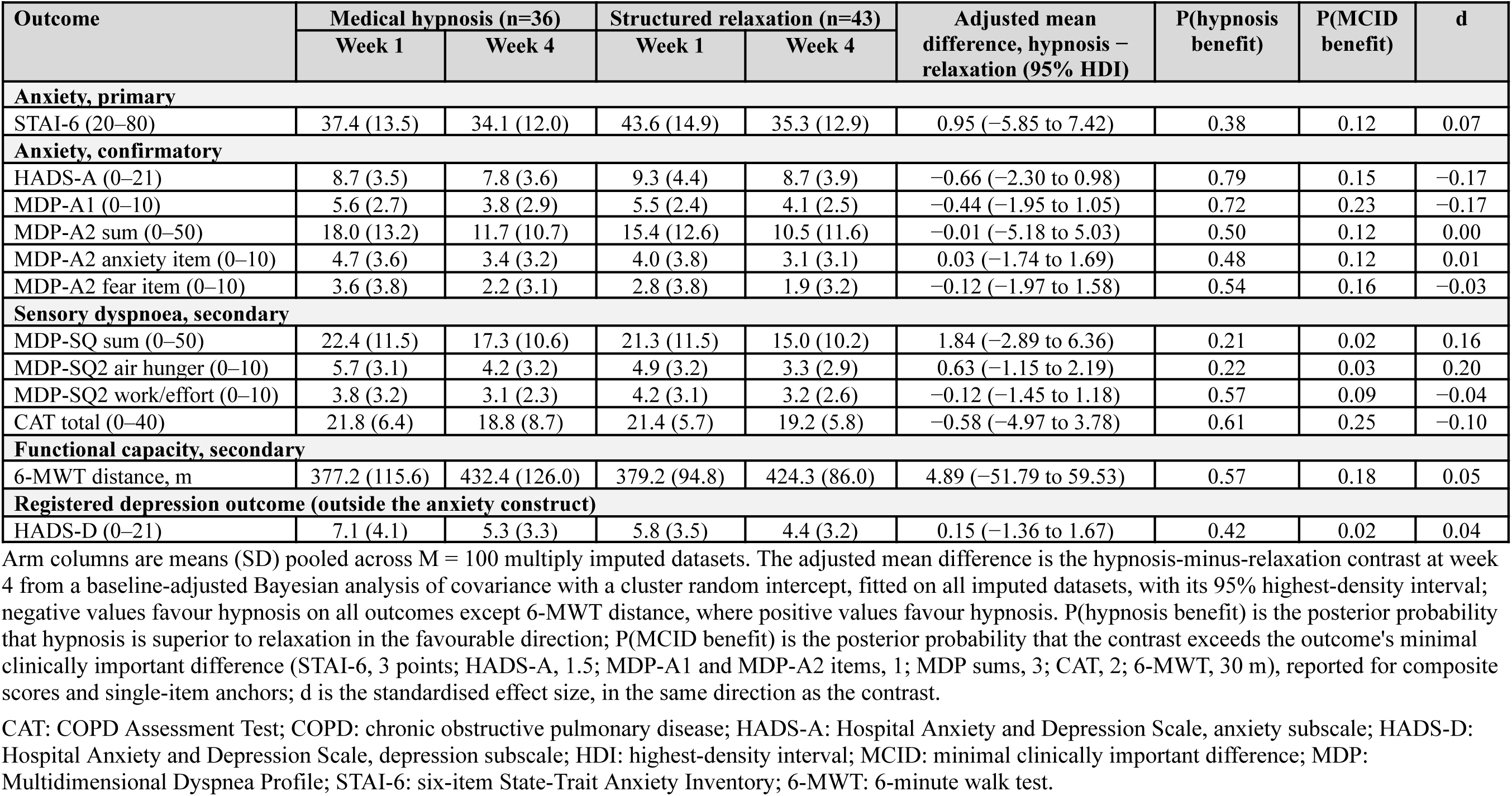
Primary and secondary outcomes at week 4 (end of pulmonary rehabilitation), pooled population

### Confirmatory and secondary outcomes

The same null pattern held across the confirmatory anxiety measures, and secondary measures of CAT and 6-minute walk test: all week-4 contrasts were centred near zero relative to their MCIDs, and the posterior probability of a clinically important hypnosis benefit was at most 0.25 (CAT) (Table 2). The intracluster correlation coefficient for week-4 STAI-6 was 0.127 (95% HDI, 0.000 to 0.439), with a design effect of 1.29 (S8.7).

### Moderation by baseline dyspnoea burden

In the post hoc moderation analysis, the hypnosis effect on STAI-6 varied with baseline burden in the hypothesised directions: the posterior probability of a positive sensory gradient (greater hypnosis benefit with higher sensory burden) was 0.91 and of a negative affective gradient 0.90 (Table 3; Figure 2A and 2B). At the high-sensory, low-affective anchor, the predicted between-arm contrast was −9.26 points (95% HDI, −23.37 to 4.71; posterior probability of exceeding the MCID, 0.81; d = −0.65). The corresponding cell of the descriptive 2 × 2 classification held 16 participants (Table S4).

**Figure 2.**
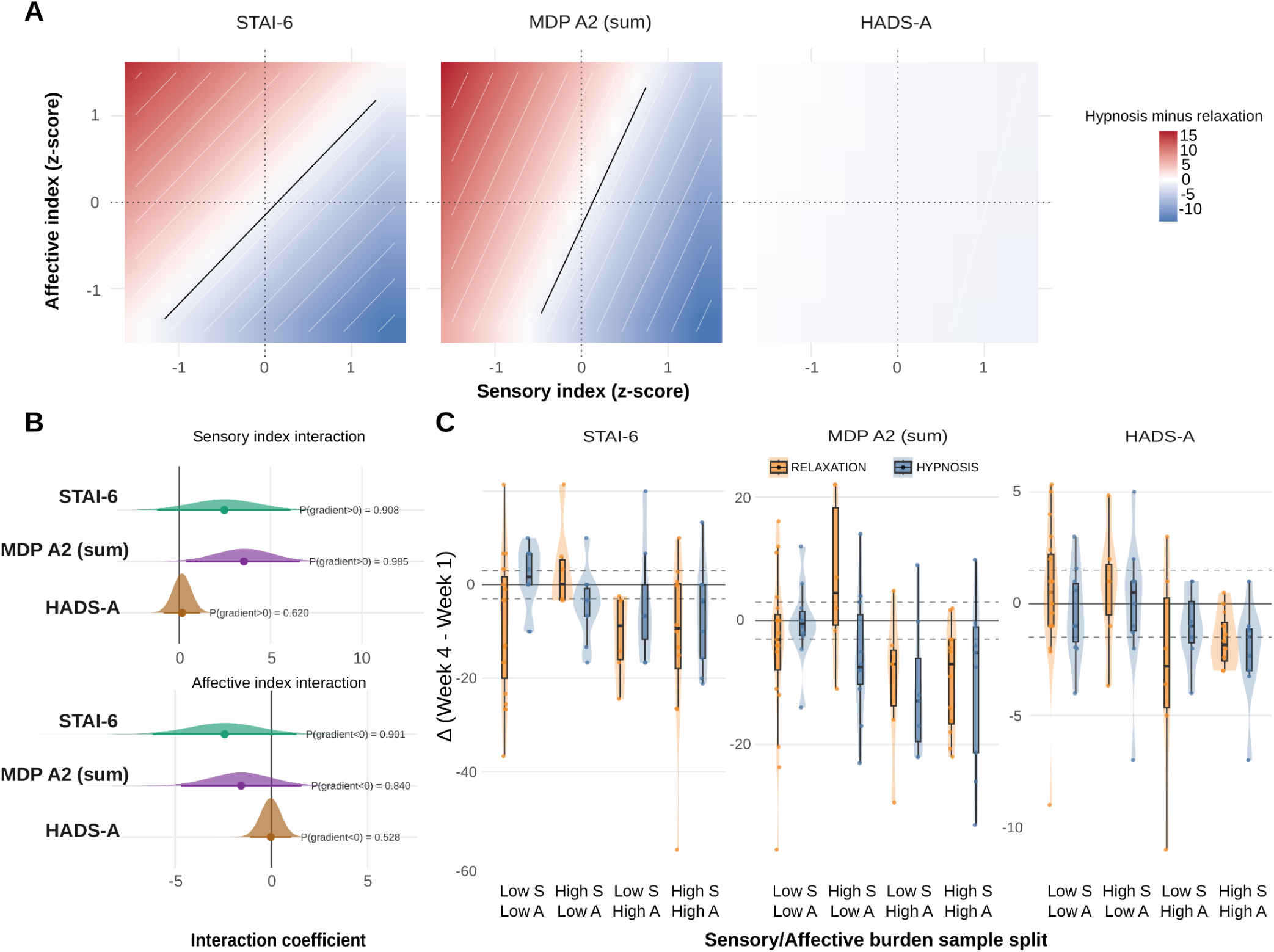
Continuous moderation of the medical hypnosis effect by baseline dyspnoea burden. (A) Posterior mean of the hypnosis-minus-relaxation baseline-adjusted week-4 contrast across the sensory × affective moderator plane, from a Bayesian analysis of covariance with continuous moderator-by-intervention interactions and a cluster random intercept (100 imputed datasets). Blue indicates predicted hypnosis benefit, red predicted relaxation benefit; the solid black line is the zero-contrast contour, and the thin white lines are contours spaced 2 points apart (±1, ±3, ±5, … points). (B) Posterior distributions of the two interaction coefficients, each annotated with the posterior probability of its hypothesised direction: for the sensory index, P(gradient > 0), the probability that hypnosis benefit increases as baseline sensory burden rises; for the affective index, P(gradient < 0), the probability that hypnosis benefit decreases as baseline affective burden rises. Under the model’s sum coding a positive gradient denotes greater hypnosis benefit as the moderator increases, consistent with Table 3. (C) Per-patient change scores (week 4 minus week 1) by baseline phenotype and arm; phenotypes are defined by splits at the mean z-score (0) on each index (low/high sensory [S] × low/high affective [A]). Dashed lines mark ±1 outcome-specific MCID (STAI-6, ±3; MDP-A2 sum, ±3; HADS-A, ±1.5); negative values indicate improvement. Y-axis ranges differ by instrument. Sensory index = mean of the standardised baseline MDP-SQ sum and CAT total; affective index = mean of the standardised baseline MDP-A2 sum, HADS-A, and STAI-6. CAT: COPD Assessment Test; HADS-A: Hospital Anxiety and Depression Scale, anxiety subscale; MCID: minimal clinically important difference; MDP: Multidimensional Dyspnea Profile; STAI-6: six-item State-Trait Anxiety Inventory.

**Table 3.** Continuous moderation of the hypnosis–relaxation contrast by baseline sensory and affective dyspnoea burden across the anxiety-construct outcomes

| Outcome | Sensory gradient, P(gradient > 0) | Affective gradient, P(gradient < 0) | Contrast at the high-sensory, low-affective anchor |  |  | P(any interaction > ½ MCID) |
|---|---|---|---|---|---|---|
|  |  |  | Predicted contrast (95% HDI) | P(MCID benefit) | d |  |
| STAI-6 (primary) | 0.91 | 0.90 | −9.26 (−23.37 to 4.71) | 0.81 | −0.65 | 0.87 |
| MDP-A2 sum | 0.985 | 0.84 | −9.17 (−20.63 to 2.40) | 0.86 | −0.73 | 0.93 |
| MDP-A2 anxiety item | 0.97 | 0.76 | −2.46 (−6.48 to 1.45) | 0.76 | −0.67 | 0.87 |
| MDP-A2 fear item | 0.93 | 0.52 | −1.63 (−5.50 to 2.27) | 0.63 | −0.43 | 0.82 |
| HADS-A (negative control) | 0.62 | 0.53 | −1.01 (−4.88 to 2.85) | 0.40 | −0.24 | 0.27 |

Post hoc, exploratory analysis. Each row is a baseline-adjusted Bayesian analysis of covariance with continuous moderator-by-intervention interactions and a cluster random intercept, fitted on M = 100 multiply imputed datasets. Sensory index = mean of the standardised baseline MDP-SQ sum and CAT total; affective index = mean of the standardised baseline MDP-A2 sum, HADS-A, and STAI-6. The anchor is +1 SD on the sensory index and −1 SD on the affective index, an illustrative location on a continuous plane rather than a subgroup.
Sign conventions: in the gradient columns the interaction coefficient is scaled so that a positive value denotes greater hypnosis benefit as the moderator increases; the sensory gradient is summarised by P(gradient > 0) (benefit increases with sensory burden) and the affective gradient by P(gradient < 0) (benefit decreases with affective burden). In the contrast column, the coefficient is the hypnosis-minus-relaxation difference on the outcome scale at the anchor, where negative values favour hypnosis (lower anxiety), as in Table 2; the standardised effect d follows the same direction. P(MCID benefit) and P(any interaction > ½ MCID) are posterior probabilities of benefit in the favourable direction.
CAT: COPD Assessment Test; COPD: chronic obstructive pulmonary disease; HADS-A: Hospital Anxiety and Depression Scale, anxiety subscale; HDI: highest-density interval; MCID: minimal clinically important difference; MDP: Multidimensional Dyspnea Profile; STAI-6: six-item State-Trait Anxiety Inventory.

The pattern was strongest and most precisely estimated on the MDP-A2 sum: sensory interaction coefficient, 3.53 per z-unit of baseline sensory burden (95% HDI, 0.36 to 6.61; posterior probability, 0.985; because the moderator is sum coded, the between-arm contrast changes by 7.06 points per z-unit), affective gradient probability 0.84, and a predicted contrast at the high-sensory, low-affective anchor of −9.17 points (95% HDI, −20.63 to 2.40; posterior probability of exceeding the MCID, 0.86; d = −0.73). Sensory gradients in the same direction were present on the MDP-A2 anxiety (0.97) and fear (0.93) items. On HADS-A, which is not anchored to the respiratory experience, neither gradient exceeded chance (0.62 and 0.53; Figure 2B).

Redefining the sensory index as the MDP-SQ sum alone left the MDP-A2 gradient essentially unchanged (0.954), whereas the STAI-6 gradient attenuated (0.908 to 0.650; Table 4). Leave-one-out affective indices and frequentist estimates of the interactions (Table S2) were concordant.

**Table 4.** Robustness of the moderation to the composition of the burden indices

| **Outcome** | **Sensory gradient, P(gradient > 0)** | **Affective gradient, P(gradient < 0)** | **Predicted contrast at the high-sensory, low-affective anchor (95% HDI)** | **P(MCID benefit)** |
|---|---|---|---|---|
| **Primary configuration: sensory index MDP-SQ sum + CAT; affective index MDP-A2 sum + HADS-A + STAI-6 (as in Table 3)** |  |  |  |  |
| STAI-6 | 0.908 | 0.901 | −9.26 (−23.37 to 4.71) | 0.81 |
| MDP-A2 sum | 0.985 | 0.840 | −9.17 (−20.63 to 2.40) | 0.86 |
| **Affective index without baseline STAI-6** |  |  |  |  |
| STAI-6 | 0.850 | 0.775 | −6.17 (−20.28 to 7.73) | 0.67 |
| MDP-A2 sum | 0.984 | 0.864 | −9.57 (−20.70 to 1.60) | 0.88 |
| **Affective index without baseline MDP-A2 sum** |  |  |  |  |
| STAI-6 | 0.915 | 0.922 | −7.91 (−20.30 to 4.38) | 0.78 |
| MDP-A2 sum | 0.980 | 0.751 | −7.32 (−18.05 to 3.54) | 0.79 |
| **Sensory index without the CAT: MDP-SQ sum alone\*** |  |  |  |  |
| STAI-6 | 0.650 | – | – | – |
| MDP-A2 sum | 0.954 | – | – | – |

Post hoc, exploratory analysis. Each row is the model of Table 3 refitted with the stated index composition and all other terms unchanged (baseline-adjusted Bayesian analysis of covariance with continuous moderator-by-intervention interactions and a cluster random intercept, M=100 multiply imputed datasets). Gradients are posterior probabilities of the hypothesised directions (greater hypnosis benefit with higher sensory burden; smaller benefit with higher affective burden). The anchor is +1 SD on the sensory index and −1 SD on the affective index; the contrast is hypnosis minus relaxation on the outcome scale, negative values favouring hypnosis, and P(MCID benefit) is the posterior probability that it exceeds 3 points in favour of hypnosis. Baseline STAI-6 and MDP-A2 sum are removed from the affective index in turn because each is also an outcome.
\*With the CAT removed, the sensitivity analysis re-estimated the sensory interaction only (posterior mean coefficient, 95% HDI: MDP-A2 sum +2.15, −0.37 to +4.61; STAI-6 +0.54, −2.29 to +3.34; the two sensory indices correlate at r=0.82); the affective gradient and the anchor contrast were not re-estimated. For comparison, the primary-configuration coefficients are +3.53 (+0.36 to +6.61) for the MDP-A2 sum and +2.46 (−1.21 to +6.09) for STAI-6.
CAT: COPD Assessment Test; HADS-A: Hospital Anxiety and Depression Scale, anxiety subscale; HDI: highest-density interval; MCID: minimal clinically important difference; MDP: Multidimensional Dyspnea Profile; MDP-A2: MDP emotional-response scale; MDP-SQ: MDP sensory-quality scale; SD: standard deviation; STAI-6: six-item State-Trait Anxiety Inventory.

### Follow-up

Telephone STAI-6 and CAT at months 1 to 3 are reported descriptively because of differential attrition (month-3 STAI-6 completion: hypnosis 50%, relaxation 84%). A tipping-point analysis required the missing hypnosis-arm month-3 STAI-6 values to be shifted by about +8 points to neutralise the week-4 advantage in the high-sensory, low-affective phenotype, and by about +13 to reverse it beyond the MCID (S8.11).

Raw between-arm contrasts remained below the MCIDs at every time point (CAT: −0.58, −0.73, and −1.79 at months 1, 2, and 3; STAI-6 in Table S6). Self-practice after discharge was modest in both arms and declined over follow-up (no practice at month 1: 64% of hypnosis and 41% of relaxation respondents). Perceived usefulness was comparable (median 5 of 7), and rehospitalisation, action-plan use, and smoking showed no consistent between-arm pattern (Table S7).

### Sensitivity analyses

Adding baseline PaCO₂ as a covariate (complete cases with this measurement available: STAI-6, n=55; MDP-A2 sum, n=54) shifted the STAI-6 contrast by −1.72 points (from 3.98 to 2.26; the starting value is the complete-case contrast within this subset, which differs from the multiply imputed full-sample estimate of 0.95) and the MDP-A2 sum contrast by 0.05 points (Table S3). Adjusting the primary model for cluster enrollment time left the contrast essentially unchanged (+1.41; 95% HDI, −5.38 to 7.80) (S8.11). Responder-rate analyses by potential phenotype were concordant with the primary results (Table S4).

### Harms

No adverse events were reported and no deaths occurred in either arm. No hypnosis-related event of interest (dissociative reaction, marked anxiety exacerbation, traumatic-memory reactivation, or distress from intrusive imagery requiring intervention) was recorded, and no participant discontinued rehabilitation because of either intervention (S10). Readmissions reported at telephone follow-up (Table S7) were collected as descriptive follow-up items, not through the adverse-event framework, and were not recorded as serious adverse events (S10).

## Discussion

PR supplemented with mind-body adjunctive care showed clinically significant decreases in anxiety at programme’s end. The adjunctive medical hypnosis arm did not differ from the adjunctive structured relaxation arm: the population-level null was uniform across the primary and confirmatory anxiety outcomes, sensory dyspnoea, and functional capacity, while anxiety improved overall by more than the MCID (pooled change, −5.72 points; S8.13). In the exploratory, post hoc moderation analysis, the hypnosis benefit on STAI-6 increased with baseline sensory burden and decreased with baseline affective burden. The sensory pattern replicated across the confirmatory anxiety measures, was strongest on the MDP-A2 sum (0.985), was absent on HADS-A (i.e. the within-construct negative control) and held on the MDP-A2 sum, under every recomposition of the burden indices. The STAI-6 gradient attenuated when recomputed without the CAT, reflecting a sensory–somatic composite rather than a purely sensory dyspnoea (Table 4). At the high-sensory, low-affective anchor, the predicted STAI-6 contrast was −9.26 points, close to three times the MCID. These results thus hint at an intervention-specific benefit moderated by baseline dyspnoea profile, and pave the way to future trials stratified on burden as a relevant diagnostic variable.

Previous trials had shown an immediate anxiolytic effect of one hypnosis session in severe inpatient COPD (9), and hypnosis attenuating both dimensions of experimentally induced dyspnoea in healthy volunteers (10). The present trial tested them in a cluster-randomised, active-comparator PR setting with Bayesian inference over multiply imputed data. Unlike waitlist- or usual-care-controlled trials (8), holding the relaxation, motivation, and contact components constant (24–26) isolated the mechanism specific to hypnosis, sensory suggestion and interoceptive redirection (11, 27, 28). The gradient analysis yielded a potential candidate responder profile: patients entering PR with high sensory and low affective dyspnoea burden may gain additional anxiety benefit from hypnosis, whereas those with high affective burden are predicted to be at least equally well served by relaxation. This is consistent with the proposal that hypnotic interoceptive redirection engages cognitive-control and attentional resources that are less available under affective dysregulation, a process-level account grounded in prior work (12, 27, 29) that our design supports, and with evidence that sensory and affective improvements in breathlessness during PR track separate brain networks (30, 31). Interestingly, the same affect-blunts-suggestion pattern has been found in gut-directed hypnotherapy for refractory irritable bowel syndrome, where higher symptom burden with lower anxiety predicted response (32), and in hypnotic analgesia, which outperforms relaxation on the sensory but not the affective dimension of chronic pain (33, 34).

For practice, the findings potentially support brief mind-body adjuncts to PR for patients with COPD and elevated anxiety, while cautioning against preferential adoption of hypnosis over relaxation for unselected patients. Anxiety improved to a clinically meaningful degree in both arms, although, in the absence of a rehabilitation-only comparator, this within-arm change cannot be attributed to the adjunctive sessions specifically. No adverse events attributable to either adjunct were recorded; both are deliverable by trained staff within existing PR resources and oriented to self-management, so continued benefit depends on uptake, modest here, which argues for structured adherence support.

In a multicentre study of 276 COPD outpatients, the affective dimension of dyspnoea was more pronounced in those with worse dyspnoea grade and symptom burden, pointing to the need of personalized treatment based on burden (35, 36). The present trial contributes to this outstanding need by presenting two different adjunctive interventions and their potential relevance for different dyspnoea profiles. Unlike trait-based clusters derived from clinical characteristics and then examined for affective load (35), burden-profile phenotypes partition patients directly on the sensory–affective plane, making the targeted dimension the classifying variable. We suggest a pragmatic next step would be a preregistered superiority trial that stratifies recruitment by baseline burden, allocates hypnosis and relaxation to the predicted candidate responder phenotypes, and tests the predicted contrast along STAI-6 MCIDs.

This trial has limitations. First, the outcomes are patient-reported, so participants were necessarily unmasked. Expectancy and credibility were balanced by randomisation and standardized via scripting, but they were not effectively measured. No suggestibility scores were obtained at any stage either. Second, despite extensive training on behalf of the practitioners, partial involuntary transfer of hypnotic communication cannot be excluded, given that these were live rather than tape-recorded interventions. Third, the comparator was embedded in a programme already delivering interoceptive components to both arms. Removing said components would have implied degrading the CHB current treatment standards, which was judged unacceptable. The present contrast captures only the increment of sensory suggestion over relaxation. Fourth, recruitment fell below target (pandemic-era reclassification, smaller cohorts). Fifth, a single trial in a French residential centre has limited generalizability, and while fidelity was assured by design (verbatim scripts, a written conduct protocol, certified practitioners rotated across arms) and monitored by a pass/fail delivery check once per cohort, we acknowledge that we did not record the sessions to monitor adherence. Sixth, the moderation was post hoc in operationalisation, not prospectively powered, and the cognitive processes invoked here are proposed interpretations based in the literature.

## Conclusions

In a PR programme that improved anxiety overall, an adjunctive medical hypnosis arm did not reduce anxiety more than a matched structured relaxation arm. The absence of a between-arm difference shows that in matched conditions the hypnotic component added nothing measurable for unselected patients. However, by indexing each patient’s sensory and affective dyspnoea burden with instruments already in routine use, this trial proposes, from an exploratory perspective, a testable candidate responder phenotype, and a framework for developing personalised, burden-based adjunctive therapy for anxiety in COPD.

## Data Availability

All data produced will be available online after publication in a peer-reviewed outlet.

## Take-Home Message

In a cluster-randomised trial in COPD pulmonary rehabilitation, medical hypnosis did not reduce anxiety more than matched relaxation. An exploratory analysis suggests greater hypnosis benefit in patients with predominantly sensory dyspnoea burden.

## Author contributions

A.G. and B.H. co-conceived the study hypothesis, led the clinical conduct of the trial and intervention fidelity, oversaw recruitment and site operations, co-wrote the manuscript, and collected all trial data. They verified data integrity and diagnostic classification. F.L. co-conceived the hypothesis and design, supervised data collection and analysis, and collected the follow-up data. D.S., as lead nurse, oversaw data collection and verified the integrity of the data-collection pipeline. I.S., C.M., and A.D. delivered the hypnosis and structured relaxation sessions and contributed to trial conduct. Y.B. contributed to data collection. T.S. and C.M.-P. contributed to data analysis and interpretation and to the implementation of the burden framework, which drew on C.M.-P.’s sensory–affective model of dyspnoea, and co-wrote the first draft. H.A. co-conceived the study hypothesis and design, developed the moderation framework operationalising baseline sensory and affective dyspnoea burden, performed the statistical analyses, interpreted the findings, co-wrote the first draft in the capacity of senior investigator, had full access to all study data after unmasking, and had final responsibility for the decision to submit for publication. All authors had access to the data, contributed to data interpretation, critically reviewed the manuscript, and approved the final version for submission.

## Conflict of interest

A.G., B.H., F.L., D.S., I.S., Y.B., C.M., A.D., T.S., C.M.-P., and H.A. reported no conflict of interest.

## Funding

This article presents independent research funded by the Agence Nationale de la Recherche (COMPSOC: ANR-24-CE28-4361; BREATHSMELLRELAX: ANR-22-CE37-0014) and the Helebor Foundation, France. The funders had no role in the design or conduct of the trial, data analysis, interpretation, manuscript preparation, or the decision to submit. The views expressed are those of the authors.

## Acknowledgements

We thank the pulmonary rehabilitation team and the patients at Centre Hospitalier de Bligny. The authors used Claude (Anthropic), an AI assistant, for formatting the manuscript and CONSORT table to the journal’s requirements, and to streamline the analysis scripts for clarity before sharing (analyses were not performed natively with AI). The authors reviewed all output, and take full responsibility for its content.

## Data availability

Anonymised individual participant data, the data dictionary, the statistical analysis code, and the intervention scripts will be deposited on the Open Science Framework at acceptance under a CC BY 4.0 licence. A repository will be shared after acceptance.

## Ethical approval

The trial was approved by the Comité de Protection des Personnes Île-de-France 1 (reference 2019-A02016-51).

## Patient consent

Written informed consent was obtained from all participants.

## Clinical trial registration

ClinicalTrials.gov NCT04868357, registered prospectively on April 29, 2021. The trial protocol has been published: Anlló H, et al. ERJ Open Res 2022;8:00565-2021 (doi:10.1183/23120541.00565-2021).

## Notes

### Competing Interest Statement

The authors have declared no competing interest.

### Clinical Trial

NCT04868357

### Clinical Protocols

https://pmc.ncbi.nlm.nih.gov/articles/PMC8819244/

### Author Declarations

The Comite de Protection des Personnes Ile-de-France 1 of Hotel-Dieu Hospital, Assistance Publique, Hopitaux de Paris gave ethical approval for this work (approval number 2019-A02016-51).

### Summary of Updates

The changes are mainly format changes that respond to a need to make this preprint accessible to a larger readership.

